# Development of a dual-gene loop-mediated isothermal amplification (LAMP) detection assay for SARS-CoV-2: A preliminary study

**DOI:** 10.1101/2020.04.08.20056986

**Authors:** Azeem Mehmood Butt, Shafiqa Siddique, Xiaoping An, Yigang Tong

**Affiliations:** Department of Biosciences, Translational Genomics Laboratory, COMSATS University Islamabad (CUI), Islamabad 45550, Pakistan; National University of Medical Sciences (NUMS), Rawalpindi, Pakistan; College of Life Science and Technology, Beijing University of Chemical Technology, Beijing 100029, China

**Keywords:** SARS-CoV-19, COVID-19, LAMP

## Abstract

Severe acute respiratory syndrome (SARS) coronavirus 2 (SARS-CoV-2) has emerged as a rapidly spreading global pathogen stressing the need for development of rapid testing protocols ever than before. The aim of present study was to develop a SARS-CoV-2 detection protocol which can be performed within minimal resources and timeframe. For this purpose, we implemented the reverse transcription loop-mediated isothermal amplification (RT-LAMP) methodology for the qualitative detection of SARS-CoV-2 RNA. In order to improve the detection capability, the RT-LAMP assay was developed to simultaneously amplify two viral genes: *ORF1a* and *N*. A total of 45 SARS-CoV-2 associated coronavirus disease 2019 (COVID-19) and 25 non-COVID-19 cases were enrolled. Viral RNA was extracted from the nasopharyngeal swab samples and analyzed simultaneously using PCR and RT-LAMP protocols. Overall, our SARS-CoV-2 dual gene RT-LAMP assay was found to be 95% accurate in detecting positive cases and showed no cross-reactivity or false-positive results in non-COVID-19 samples. Further evaluation on larger and multi-centric cohorts is currently underway to establish the diagnostic accuracy and subsequent implementation into clinical practice and at point-of-care settings.

## INTRODUCTION

Severe acute respiratory syndrome (SARS) coronavirus 2 (SARS-CoV-2) is an enveloped, non-segmented single-stranded, positive-sense RNA virus comprising of 29,881 nucleotides in length. At genomic level, SARS-CoV-2 shares ∼88% nucleotide sequence homology with previously reported SARS-like coronavirus strains from bats, indicating its potential origin and transmission route [1].

SARS-CoV-2 associated coronavirus disease 2019 (COVID-19) has rapidly become a global public health emergency since its outbreak in Wuhan, China in December 2019 [2]. The current recommended testing method for patients exhibiting clinical features of COVID-19 is the qualitative detection of SARS-CoV-2 via real-time reverse transcription polymerase chain reaction (rRT-PCR). The rRT-PCR for SARS-CoV-2 can be performed on nasopharyngeal and/or oropharyngeal swabs, sputum or alveolar lavage fluid depending upon the sample processing local/international guidelines and available facilities [3, 4]. However, COVID-19 outbreak has highlighted several caveats of the PCR-based testing when it comes down to the point of mass-testing coupled with urgent reporting. For instance, PCR testing requires well-equipped centralized laboratories capable of handling respiratory pathogens, dedicated instruments such as PCR machines and skilled staff to ensure the smooth processing which isn’t always possible particularly in less developed or resource limited communities. Unfortunately, the COVID-19 has pushed even the developed nations to levels where they are struggling to ensure rapid and effective testing for every suspected case, one can imagine how catastrophic the situation can be in countries with limited healthcare facilities and resources. In addition, false-negative PCR results from several COVID-19 patients is also becoming a matter of concern raising questions about the accuracy of SARS-CoV-2 PCR assays [5-7]. Therefore, a great deal of effort is shifting towards deployment of accurate yet rapid, cost-effective and point-of-care testing methodologies.

As the basic principle of PCR-based methods is to amplify the genomic content to the level of detection, alternative molecular amplification approaches can be of great value under crisis situations. One such method is loop-mediated isothermal amplification (LAMP) which since its development became one of the highly popular and implemented isothermal method for the rapid detection of animal [8, 9], plant [10, 11] and human pathogens [12, 13]. Unlike PCR, LAMP utilizes a combination of uniquely designed pair of four to six primers and a specialized version of DNA polymerase with strand displacement activity to bypass the need of denaturation by heat. In summary, it is well-established that LAMP is a stable and rapidly deployable approach with sensitivity profile surpassing the PCR [10, 14, 15]. Recently, several LAMP assays for the detection of SARS-CoV-2 have been reported and showed promising data [16-23]. In the present study, we report a novel SARS-CoV-2 dual gene RT-LAMP assay and its preliminary evaluation on “real-world” cases.

## MATERIAL & METHODS

### Study design

The present study aimed towards development and assessment of a rapid SARS-CoV-2 detection protocol based upon the concept of LAMP. Informed consent from study participants and ethical approval was obtained as per institutional guidelines.

### Study cohort

A total of 45 PCR-confirmed positive cases of COVID-19 and 25 non-COVID-19 cases were selected for the present study. The steps of rRT-PCR-based detection of COVID-19 infection were conducted as follows. All the procedures were performed in a biosafety level-3 facility as per instructions set forth for the handling of COVID-19 samples.

#### Viral RNA isolation

Total Viral RNA was extracted from the deactivated NP swab samples by using viral RNA kit following manufacturer’s recommendations. The eluted viral RNA was stored at −80°C until further use.

#### cDNA synthesis

The eluted viral RNA was reverse transcribed by using Hifair II 1^st^ stand cDNA Supermix (Yeasen, China) as per manufacturer’s recommendations. Briefly, 10ul of Supermix containing optimized concentration of Hifair II reverse transcriptase, RNase inhibitor, dNTPs, random hexamer/oligo dT primer mix in an optimized buffer system was mixed with 5ul of Viral RNA followed by addition of 5ul DNase/RNase free water (Invitrogen, USA) for a final reaction volume of 20ul. The procedure was performed on MiniAmp thermocycler (Applied biosystems, USA) and the cycling conditions were 25°C for 5min followed by 42°C for 30min and final elongation/termination at 85°C for 5min.

#### COVID-19 rRT-PCR assay

The COVID-19 rRT-PCR was performed using 2X EasyTaq PCR Supermix (TransGen, China) and China CDC approved primer (Forward primer: CCCTGTGGGTTTTACACTTAA; Reverse primer: ACGATTGTGCATCAGCTGA) and probe (5’-FAM-CCGTCTGCGGTATGTGGAAAGGTTATGG-TRAMA-3’) pair targeting *ORF1ab* gene segment. Briefly, 1ul of cDNA was added to the mix comprising of 10X EasyTaq buffer (2ul), 2.5nM dNTPs (1.6ul), EasyTaq DNA polymerase (0.3ul), 10uM primer forward/reverse (0.4 ul) 10uM probe (0.2ul), and nuclease free water (14.5ul) to makeup a final reaction volume of 20ul. The amplification was performed on QuantStudio qPCR system (Applied biosystems, USA) and the cycling conditions were; 50°C for 2min followed by denaturation step at 95°C for 10min and 40 cycles of 95°C for 10s and 60°C for 1min.

#### LAMP assay development

The full-length genome sequences of SARS-CoV-2 were downloaded from GISAID [24] (accessed on March 10, 2020). The multiple sequence alignment was performed using MAFFT [25] with manual correction of alignment files in alignment editor wherever necessary. The consensus SARS-CoV-2 genome sequence as representative of multiple sequence alignment was generated for further analysis. The LAMP primers were designed using Primer Explorer v5 (http://primerexplorer.jp/lampv5e/index.html) software targeting the *ORF1a* and *N* gene segments of consensus genome sequence. Multiple primer pairs were generated which were further evaluated based upon principles of LAMP primer design. Each primer set comprised of an outer forward primer, outer backward primer, forward inner primer, backward inner primer, loop forward primer, and loop backward primer. The pair of primers (available upon request) targeting *ORF1a* and *N* genes that were found to be non-homologous to non-COVID-19 coronavirus strains based upon BLAST analysis.

The RT-LAMP reaction was performed using WarmStart Colorimetric LAMP 2XLJMaster Mix (New England Biolabs, Beverly, MA, United States). Briefly, 10ul of WarmStart master mix was mixed with 2ul of 10x primer mix, 3ul of template viral RNA and adjusted with 5ul of water for a final volume of 20ul. The reaction tubes were incubated at 65°C for 5min in a pre-heated dry bath. Blank and negative controls were included in each cycle. Results were interpreted based on visualization of change in color once the amplification was terminated. Positive samples turned yellow whereas negative samples remained pink.

### Statistical analysis

The Fisher’s exact test was used to computer the diagnostic accuracy of dual-gene RT-LAMP assay in comparison to rRT-PCR.

## RESULTS & DISCUSSION

In the present study, we report a novel dual-gene RT-LAMP assay for the qualitative detection of SARS-CoV-2 in COVID-19 cases. As the aim is to mature this protocol as a point-of-care and on-field testing approach, therefore, the protocol was kept simple, quick and easily deployable. For this purpose, ready-to-use master mixes and primer mixes were prepared. Multi-gene approach has previously shown to improve the performance of SARS-CoV-2 PCR assays. The same approach was applied here, and RT-LAMP was designed to simultaneously detect *ORF1a* and *N* genes. Recently, several studies have reported LAMP assays for COVID-19. Overall, all these studies highlight the strength of LAMP especially in situations where a rapid and reliable testing is required. Moreover, few commercially developed isothermal systems are already in pipeline for the approval by the relevant authorities however, majority of the reported SARS-CoV-2 LAMP assays were evaluated on synthetic constructs and lack validation on actual samples. Theoretically, such assays shouldn’t suffer from any drastic differences, however, the true sensitivity: specificity profile of an assay demands validation on clinical cohort. Therefore, one strength of our assay is validation on COVID-19 positive patients’ samples. As given in Table 1, dual-gene LAMP assay was able to detect 43 out of 45 PCR-confirmed SARS-CoV2 positive samples based upon consensus findings of *ORF1a* and *N* genes whereas, none of the non-COVID-19 samples were found to be positive indicating 95% sensitivity and 100% specificity.

**Table 1.** Comparison of diagnostic accuracy of dual gene RT-LAMP vs. PCR.

| Dual gene RT-LAMP assay | rRT-PCR assay |  | Total |
| --- | --- | --- | --- |
|  | Positive (n) | Negative (n) |  |
| Positive (n) | 43<br>(*True Positive*) | 0<br>(*False Positive*) | 43 |
| Negative (n) | 2<br>(*False Negative*) | 25<br>(*True Negative*) | 27 |
| Total (n) | 45 | 25 | 70 |
| Sensitivity (%) | 95 |  |  |
| Specificity (%) | 100 |  |  |
| Positive Predictive Value (PPV (%)) | 100 |  |  |
| Negative Predictive Value (NPV) (%) | 95 |  |  |

The two PCR-positive samples designated as sample A (ID#1112-4) and B (ID# 390-3) showed conflicting results. As shown in Figure 1, sample A showed positive amplification in *N* assay whereas no amplification was noted with *ORF1a* assay. On the other hand, sample B showed positive amplification with *ORF1a* whereas negative according to the *N* assay. As we considered same output from both assays to assign samples as positive or negative, we concluded these samples as negative. The Cq value of sample B was 36.90 that is indicate of low viral load whereas, sample A had Cq value 31.30. As there was no additional viral RNA available for testing of sample A and B, the negative result of sample A for *ORF1a* is currently unclear. On the other hand, positive detection of low viral load sample B is indicative that LAMP *ORF1a* assay is more sensitive than *N* assay. Evaluation with additional samples is currently underway. Overall, the reported dual-gene LAMP assay is an effective and quick approach to handle mass-testing of COVID-19 without the need of expensive equipment, access to centralized laboratories and highly trained staff.

**Figure 1.**
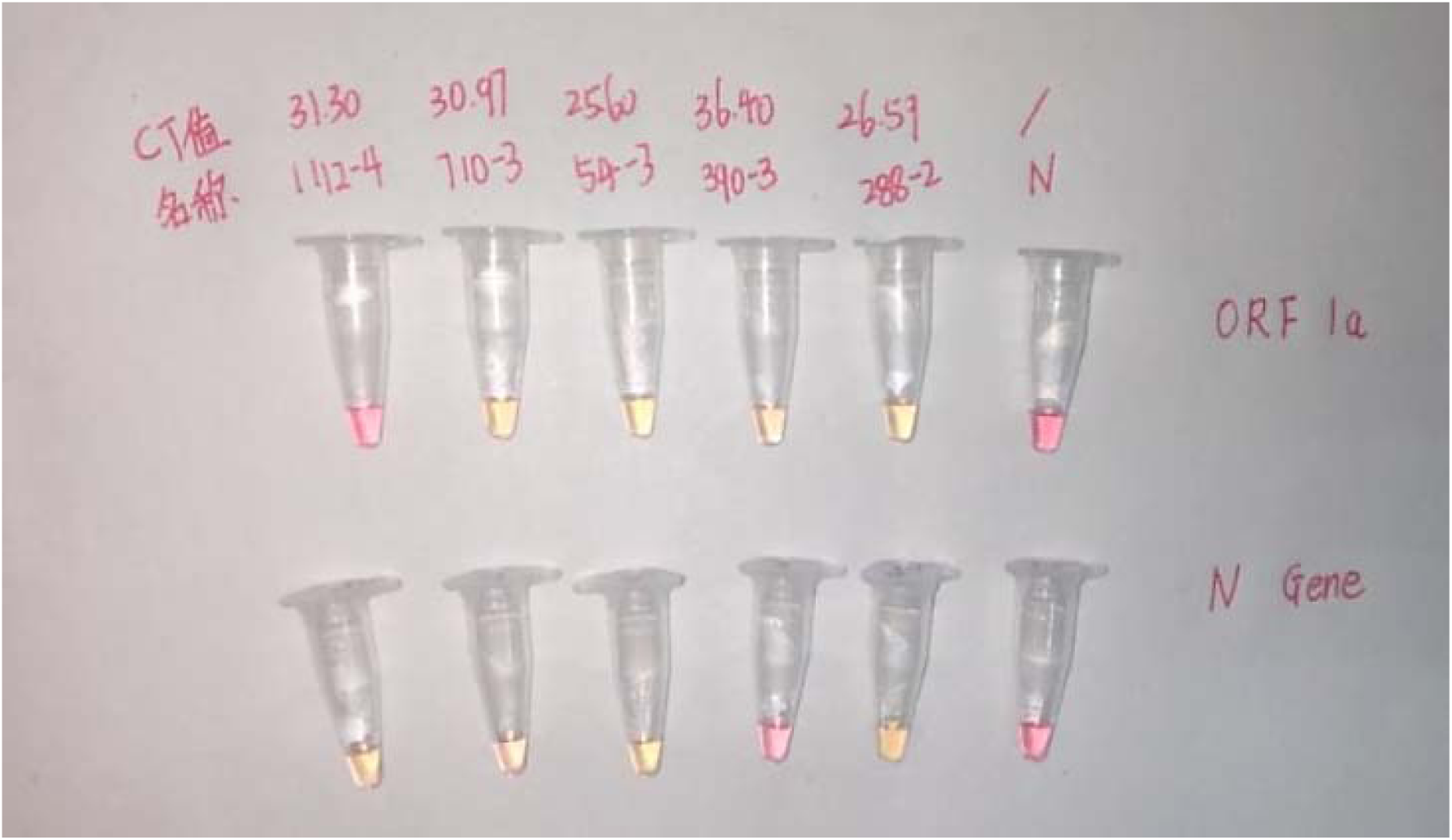
Comparative analysis of dual gene LAMP vs. PCR.

The representative image shows randomly selected samples from study cohortwith findings in agreement to rRT-PCR and two samples that had conflicting results sample A (ID#1112-4) and sample B (ID# 390-3). The top row refers to the Cq values. Middle row refers to sample IDs. N; Negative control.

## Data Availability

Data is provided into the manuscript.

## Acknowledgements

Acknowledgments are to clinicians, study participants and hospital staff for their assistance in samples and data collection.

